# A Reproducible Pipeline for Processing Commercial Wearable Step-Count Data in Aging Cohorts: Application and Evaluation in the STRRIDE-PD Reunion Study

**DOI:** 10.64898/2026.05.14.26353213

**Authors:** Nannan Bo, Alyssa M. Sudnick, Julie D. Counts, Katie G. Kennedy, Agustin A. Saldana, Katherine A. Collins-Bennett, William C. Bennett, Johanna L. Johnson, Kim M. Huffman, Amanda E. Paluch, Marissa C. Ashner, William E. Kraus, Sarah B. Peskoe, Leanna M. Ross

## Abstract

Wearable devices offer the ability to objectively characterize free-living physical activity; however, raw step-count data generated by commercial devices require systematic processing before they can support rigorous inference. We describe a transparent, reproducible standard operating procedure (SOP) for transforming epoch-level step-count data from commercial Garmin devices into participant-level analytic variables and demonstrate its application in the STRRIDE-PD Reunion study: a long-term follow-up of older adults originally enrolled in a supervised exercise intervention trial. This data pipeline standardizes timestamps, reconstructs daily epoch grids, infers wear time from observed step patterns, and applies a prespecified valid-day threshold (≥10 hours inferred wear time) to generate participant-level summaries. Among 67 participants (mean age 71.4 years, 65.7% women), the median valid-day count was 10 days, median average daily steps were 5,794, and participant-level estimates were identical across ≥10-hour and ≥6-hour valid-day thresholds. Wearable-derived step counts were significantly associated with 9 of 16 cardiometabolic and fitness outcomes, including cardiorespiratory fitness, body composition, and lipid profiles. By contrast, self-reported exercise – assessed via a frequency-by-duration composite ranked into deciles – was not significantly associated with any outcome. A regression calibration framework applied to the full sample quantified the attenuation underlying this discrepancy: the naive self-report model systematically underestimated associations relative to both the observed Garmin model and calibration-corrected estimates. These findings demonstrate that measurement approach is a determinant of scientific conclusions in physical activity research, and that reproducible wearable data pipelines are essential infrastructure for aging epidemiology.

**Highlights:**

- A reproducible standard operating procedure processed Garmin step-count data without wear-time indicators.
- Wearable steps predicted 9 of 16 outcomes; self-reported exercise predicted none.
- Regression calibration revealed that self-report systematically underestimated physical activity-health associations.
- Measurement approach determines physical activity-health conclusions in aging cohort research.

## 1. Introduction

Physical activity (PA) is a key determinant of cardiovascular health, metabolic disease, and premature mortality across the life course (Lee and others 2012). Higher levels of habitual PA are associated with preserved cardiometabolic health, reduced risk of type 2 diabetes, and delayed functional decline (Aune and others 2015; Booth and others 2012; Pahor and others 2014; Paterson and Warburton 2010; Piercy and others 2018). These associations are particularly pronounced in older adults with even modest increases in PA reducing the risk of major mobility disability and functional limitation by 40–50% in community-dwelling individuals aged 65 and older (Pahor and others 2014; Paterson and Warburton 2010). As a result, accurate and scalable measurement of free-living PA is critical for epidemiologic and clinical research, particularly in studies seeking to understand long-term activity patterns and their relationships with health outcomes.

Historically, in large epidemiologic and clinical studies, PA has been assessed primarily through self-reported questionnaires (Prince and others 2008). Decades of large-scale epidemiological research have provided the empirical foundation for much of what is known about PA and chronic disease risk (Arem and others 2015). While inexpensive and easy to administer, self-reported PA measures are prone to recall bias, social desirability bias, and misclassification, often showing limited agreement with objective measures of activity (Adams and others 2005; Dyrstad and others 2014; Prince and others 2020). Social desirability pressure causes systematic overreporting of PA on self-report instruments (Adams and others 2005), and agreement between self-report and accelerometry is particularly weak in older adults (Dyrstad and others 2014). Moreover, questionnaire-based PA measures typically capture only intentional, structured exercise sessions and miss incidental walking, active transportation, and other habitual daily movement which constitutes the majority of activity-related energy expenditure.

Within the past 10-15 years, consumer wearable devices (e.g. commercially available fitness trackers and smartwatches, such as Garmin) have become sufficiently ubiquitous and accurate to support their deployment in large cohort studies (Evenson and others 2015; Evenson and Spade 2020; Fuller and others 2020), enhancing opportunities to objectively assess free-living PA (Patten and others 2026). In particular, step counts derived from wearable devices provide an intuitive and scalable summary of overall ambulatory activity. Despite the advantages of consumer wearable devices, step-count data derived from these devices require several analytic decisions before the data can be used in research. In practice, this process typically involves defining valid observation days, handling periods of missing or implausible step counts, and summarizing activity across days to generate participant-level metrics such as mean daily steps (Migueles and others 2017). These analytic decisions can vary across studies depending on device characteristics, availability of metadata (*i.e.,* wear-time indicators), and practical constraints of real-world data collection, underscoring the need for transparent and reproducible processing standards.

The need for transparent and reproducible processing approaches arose in the STRRIDE-PD Reunion study, a long-term follow-up of participants originally enrolled in a supervised exercise intervention trial. During the parent STRRIDE-PD intervention phase, exercise exposure was well prescribed and monitored, and PA was assessed using self-report measures. Approximately a decade later, the Reunion study aimed to additionally characterize participants’ current PA under free-living conditions. Therefore, wearable step-count data provided an opportunity to objectively assess current PA, beyond self-report, in an aging population with well-characterized cardiometabolic and fitness outcomes, enabling direct evaluation of PA–health associations.

PA data derived from commercial wearable devices require systematic processing to distinguish plausible activity from potential non-wear, as these devices lack explicit wear-time indicators. To address this need, we developed an open-source standard operating procedure (SOP) for processing step-count data that makes analytic decisions explicit and reproducible. The pipeline-derived objective PA data also created the unique opportunity to directly evaluate how much the choice of measurement instrument – self reported versus device-measured PA – shapes scientific conclusions. We describe the SOP, demonstrate its application in the STRRIDE-PD Reunion study, evaluate the associations between pipeline-derived step counts and health-related outcomes, and apply a regression calibration framework to formally quantify the inferential consequences of using self-reported rather than device-measured PA.”

## 2. Methods

### 2.1 Study Design and Data Collection

The parent STRRIDE-PD randomized trial (1R01DK081559; NCT00962962; 2009-2014) evaluated the effects of different amounts and intensities of aerobic exercise – with and without diet-induced weight loss – on cardiometabolic health outcomes in previously sedentary adults (45-75 years) with overweight or obesity and prediabetes (Slentz and others 2016). Approximately 10 years after STRRIDE-PD completion, participants were invited to return for observational follow-up as part of the STRRIDE-PD Reunion study (R21AG075379; 2019-2024), which assessed the sustained effects of previous structured exercise training on cardiorespiratory fitness, body composition, and cardiometabolic risk factors. In the Reunion phase, PA was assessed via self-report and with wearable devices. A brief description of the health-related outcome assessments is provided in **Supplemental Table S1**. The study protocol was approved by the Duke University Health System Institutional Review Board (Pro00014088). All participants provided written informed consent.

Self-reported frequency and duration of recent PA were assessed using two questionnaire items: 1) “In the past 3 months, how frequently did you exercise?” with response options including: no exercise; once, twice, three times, or four or more times per week; and 2) “Approximately how much time did you spend exercising for each session?” with response options including: 0–20, 20–30, 30–40, 40–50, 50–60, or 60+ minutes. A composite weekly exercise volume was derived by multiplying reported frequency by the midpoint of the reported duration category. Because both items are categorical and duration categories are unequally spaced, this composite was ranked into deciles across participants (range 0-9), with higher deciles reflecting greater self-reported weekly exercise volume. Participants reporting no exercise were assigned a decile of 0.

Step-count data were obtained from commercially available Garmin vívosmart^®^ 5 devices. Participants were asked to wear the device 24 hours per day (except for brief removal for charging) for at least seven days. Data retrieval was facilitated by the Garmin Connect and Pattern Health applications (Garmin Connect: https://connect.garmin.com; Pattern Health: https://pattern.health). Garmin devices aggregated step-count data in 15-minute epochs, retrieved via the Pattern Health platform. Raw data included a time zone offset variable, which was applied to standardize epoch timestamps during data consolidation. The SOP described in Section 2.2 was then applied to transform these raw data into participant-level step-count variables.

### 2.2 Standard Operating Procedure for Step Count Data

Our standard operating procedure (SOP) described a structured workflow to transform raw Garmin device data into cleaned participant-level step-count variables suitable for statistical analysis. The primary objective was to ensure transparency and reproducibility across all stages of data processing, from raw interval-level records to analytic summaries.

Raw consumer wearable data typically consist of timestamped step-count values recorded at fixed intervals, known as epochs. However, these data often contain irregular time intervals, missing segments, and device-specific formatting inconsistencies. The proposed workflow addressed these challenges by standardizing timestamps, reconstructing daily epoch grids, explicitly inferring wear time based on observed step patterns, and generating daily and participant-level summaries using prespecified criteria for valid days.

#### 2.2.1 Key Operational Definitions

An **epoch** is the smallest unit of time over which step counts are recorded. In the present application, Garmin devices aggregated step-count data in fixed 15-minute intervals. All processing was conducted at the epoch level prior to daily aggregation.

**Wear time** is conceptually defined as the continuous period during which a participant wears the device and can generate valid step-count data. Because direct wear-time indicators (e.g., an on-body detection sensor or device-generated wear-log) were not available, wear time was operationally inferred from observed step patterns. Within each calendar day, the wear window was defined as the interval between the first and last detected non-zero step epochs. Zero-step epochs occurring within this window were retained and treated as potential inactivity. This conservative definition was chosen deliberately; participants were instructed to charge devices only during brief non-wear periods such as showering, so missed steps during charging were expected to be minimal, and anchoring the wear window to observed steps was preferred to err on the side of including only well-observed time.

A calendar day was classified as a valid wear day if at least 10 hours of inferred wear time were observed. This threshold is consistent with widely adopted conventions in accelerometer-based PA research, having been codified in national surveillance programs such as the NHANES and validated in systematic reviews of accelerometer processing criteria (Choi and others 2011; Migueles and others 2017; Tudor-Locke and others 2012). Participants were required to contribute at least four valid days to be included in the analytic sample, consistent with evidence that four days of pedometer data reliably estimate habitual PA in older adults (Hart and others 2011). All daily summaries were generated a) including all observed days and b) restricted to valid days to permit sensitivity analyses.

#### 2.2.2 Data Processing Pipeline

Raw epoch-level step-count data were processed according to the prespecified workflow illustrated in **Figure 1**. First, raw data were imported and inspected for structural consistency, timestamp formatting, and completeness. Timestamps were standardized to a common format, and duplicate or irregular records were removed. Step counts were then mapped to a complete sequence of fixed 15-minute epochs for each calendar day, with missing epochs within the observed time span retained to preserve temporal continuity.

**Figure 1.**
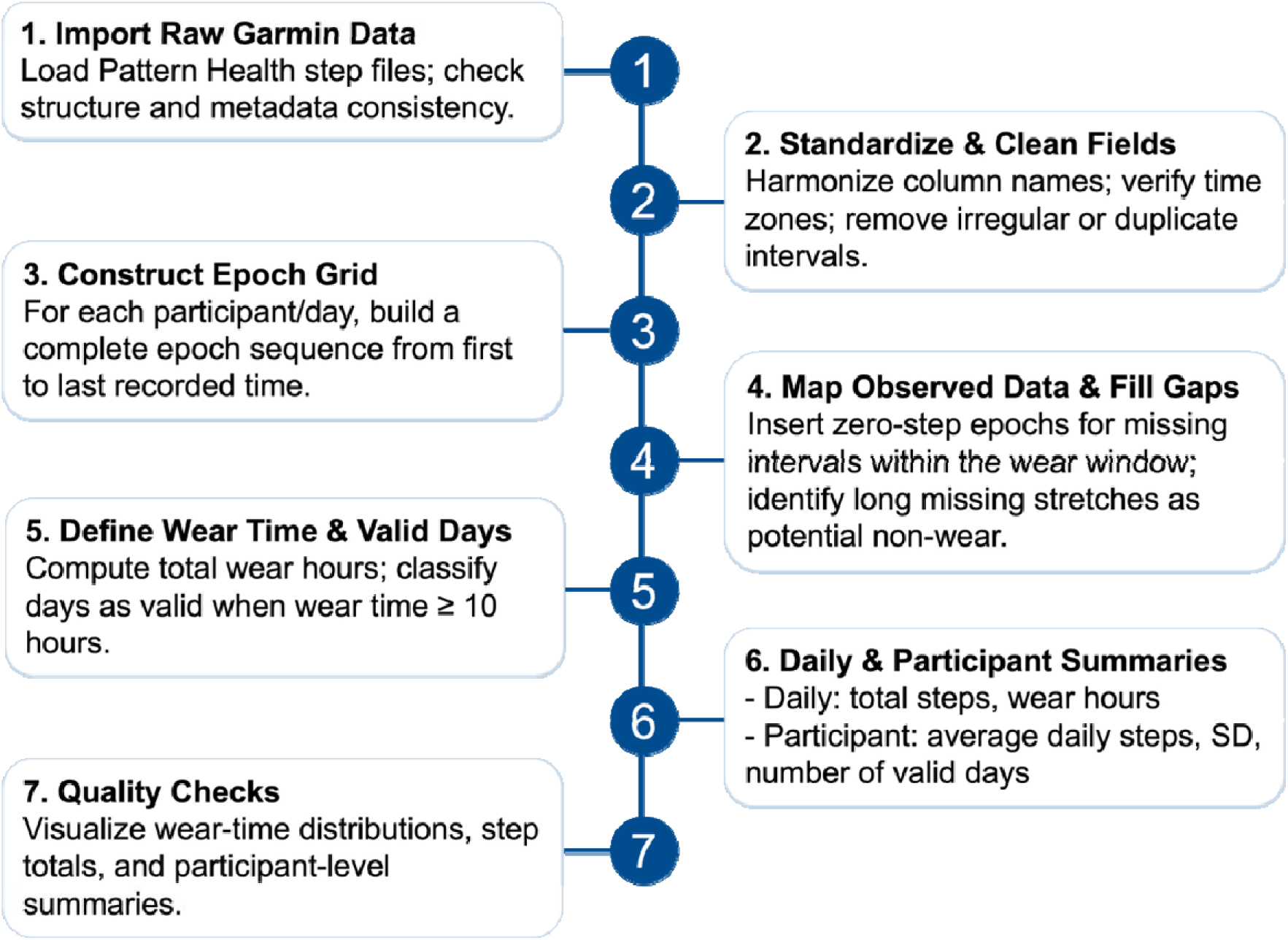
Standard operating procedure workflow

Wear time was inferred from observed step patterns, with the interval between the first and last non-zero step epochs within a day defining the wear window. Missing epochs within this window were treated as potential inactivity and assigned zero steps for aggregation. Days with at least 10 hours of inferred wear time were classified as valid and included in primary summaries; total wear time and daily steps were computed for all days to support sensitivity analyses. Participant-level metrics (including mean daily steps, within-person variability, and number of valid days) were derived from the valid-day set for downstream analyses.

Quality control procedures were applied at each stage of the pipeline to detect irregular step-count patterns that could reflect device malfunction, data corruption, or substantial non-wear. Specific checks included: flagging epochs with biologically implausible step counts, identifying irregular recording intervals, and reviewing participant-level distributions of daily wear time and daily step totals via histograms and boxplots prior to analytic aggregation. In the present dataset, one irregular recording interval of 300 minutes was identified on a single participant-day during quality control; as the cause could not be determined from the data alone, this epoch was excluded from wear-time calculations and step aggregation for that day. No participants were excluded based on data quality. The operational definitions embedded within this SOP (the 10-hour wear-time threshold and 4-day minimum) represent justified, context-specific choices rather than universal standards. The modular pipeline structure allowed investigators to modify key parameters without altering the underlying workflow.

To characterize the analytic sample, summary statistics (median, range, mean, and SD where applicable) were calculated for valid-day counts and participant-level step-count metrics, including mean daily steps, within-person SD, and coefficient of variation (CV), overall and separately under the ≥10-hour and ≥6-hour valid-day thresholds. Threshold-specific summaries were compared to evaluate the sensitivity of participant-level estimates to the choice of valid-day definition.

The complete processing pipeline and analysis code are publicly available at: https://github.com/nannanbo/stepcount-pipeline.

### 2.3 Evaluation of Self-Reported Physical Activity Using Regression Calibration

To formally evaluate the degree to which self-reported exercise may bias associations with the health-related outcomes assessed at the Reunion timepoint, we applied a regression calibration framework (Carroll and Carroll 2006; Keogh and others 2020; Rosner and others 1989). Regression calibration proceeds in two stages. In the first stage, a calibration model is fit to estimate the relationship between the error-prone measure and an objective reference, conditional on covariates. In the second stage, the calibration-predicted values (the model-fitted estimates of the objective measure for each participant) are substituted for the error-prone measure in the outcome models of interest. This substitution recovers associations that measurement noise would otherwise suppress. Here, the participants with both self-reported and Garmin data served as an internal calibration sample.

Formally, let x denote the Garmin mean daily steps (the objective reference), *x*\* denote the error-prone self-reported exercise decile, and z denote a vector of covariates (age, sex, treatment group). The calibration model regressed log *x* on *x\** and z:

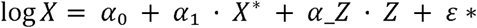

The calibration-predicted values Ê[log *x* |*x*\*, *z*] were then substituted for x* in linear outcome models for each of 16 reunion biometric outcomes. Bootstrap standard errors (B = 2,000) were used to propagate calibration uncertainty through to the corrected estimates, as recommended by Keogh and others (2020).

To provide interpretive context for the RC-corrected estimates, results are presented as a three-model comparison for each outcome: (1) a naive model using the self-reported exercise decile directly; (2) a reference model using observed log-transformed Garmin steps; and (3) the RC-corrected model using calibration-predicted values. This structure makes the direction and magnitude of bias directly visible and allows the RC-corrected estimate to be interpreted relative to both the uncorrected self-report and the objective reference. Full technical details, including model assumptions, diagnostics, and the log-transformation decision for the reference measure, are provided in Supplement S1.

## 3. Results

### 3.1 Step Count Data Quality and Summary

At the Reunion timepoint, the analytic sample comprised 68 participants (71.6 ± 7.2 years; 64.2% women) with both wearable and self-report data available. Participant characteristics are summarized in **Table 1**. One participant was excluded for contributing fewer than four valid days under the ≥10-hour wear-time threshold.

**Table 1.** Participant characteristics (n = 67)

| Variable | Summary |
| --- | --- |
| *Demographics* |  |
| Age (years), mean (SD) | 71.4 (7.3) |
| Sex, n (%) |  |
| Women | 44 (65.7%) |
| Men | 23 (34.3%) |
| Race, n (%) |  |
| Asian | 2 (3.0%) |
| Black | 11 (16.4%) |
| White | 53 (79.1%) |
| *Anthropometric, mean (SD)* |  |
| BMI | 29.1 (3.5) |
| *Original Intervention Group, n (%)* |  |
| High/Mod | 16 (23.9%) |
| High/Vig | 22 (31.3%) |
| Low/Mod | 12 (19.4%) |
| Low/Mod/Diet | 17 (25.4%) |
| *Cardiorespiratory fitness, mean (SD)* |  |
| Absolute peak $\dot{V}\text{O}_2$ (L/min) | 1.8 (0.6) |
| Relative peak $\dot{V}\text{O}_2$ (mL/kg/min) | 21.9 (5.6) |
| *Physical activity measures, median (Q1, Q3)* |  |
| Mean daily steps per individual | 5,794 (4,162, 7,989) |
| Exercise volume decile | 5.0 (2.5, 7.0) |

Note. Continuous variables reported as mean (SD); categorical variables as n (%). High/Mod indicates high-amount/moderate-intensity aerobic training; High/Vig, high-amount/vigorous-intensity aerobic training; Low/Mod, low-amount/moderate-intensity aerobic exercise; Low/Mod/Diet, Low/Mod exercise plus diet-induced weight loss.

Among the 67 participants with at least four valid days under the ≥10-hour threshold, the median number of valid days per participant was 10 (range: 6-40) (**Table 2**). Although participants were instructed to wear the device for at least seven days, many continued wearing it beyond the instructed window; 38 participants contributed more than 10 valid days. On valid days, daily wear time was concentrated between 15 to 20 hours, consistent with near-full-day device use in most participants. Across the analytic sample, mean daily steps ranged from approximately 1,000 to 15,000, with a median of 5,794 (IQR: 4,162-7,989) steps/day. The distribution was right-skewed, with a small number of participants accumulating substantially more steps than the median (**Figure 2**). The mean within-person CV was 0.355, indicating moderate day-to-day variability in step counts.

**Table 2.**
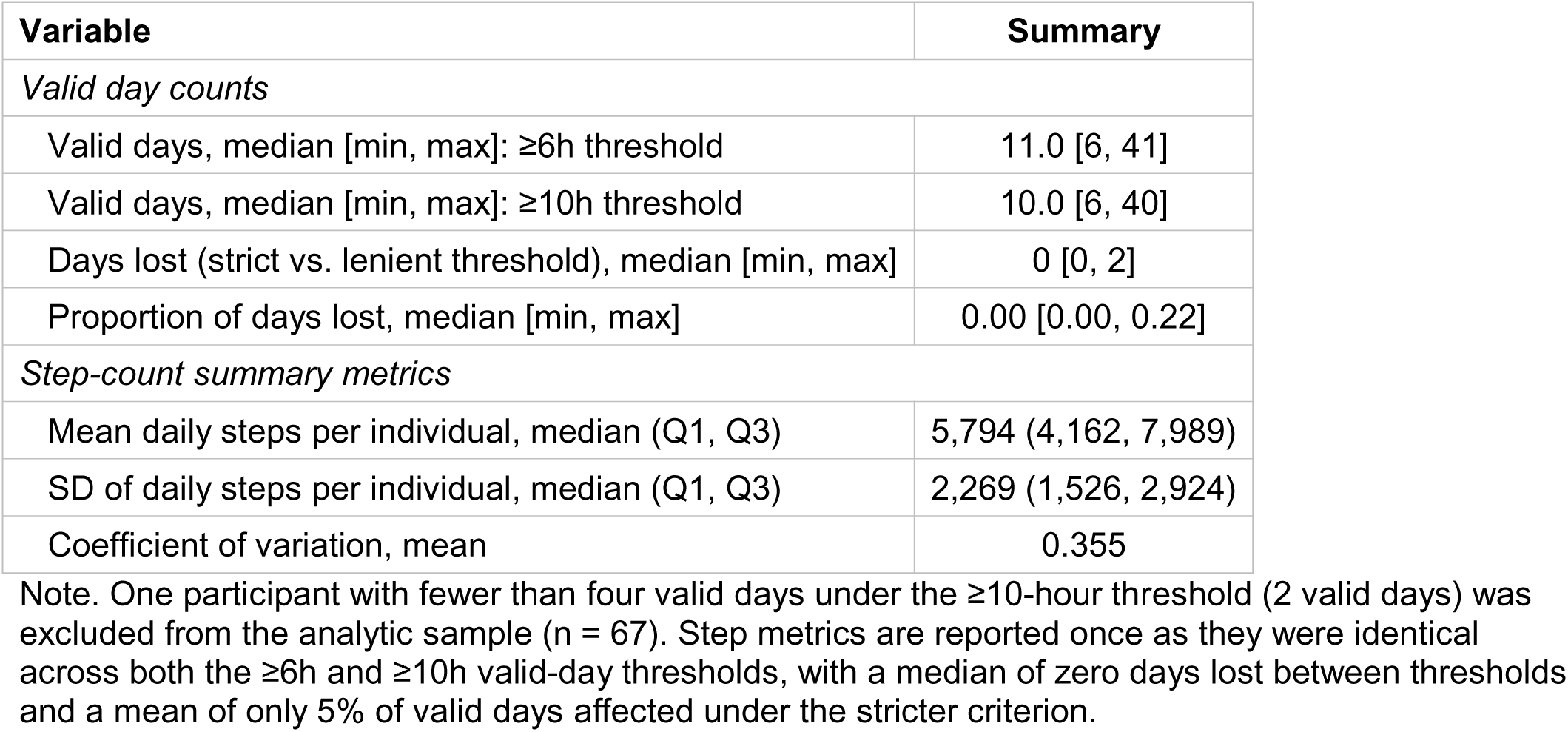
Wearable data quality summary (n = 67)

**Figure 2.**
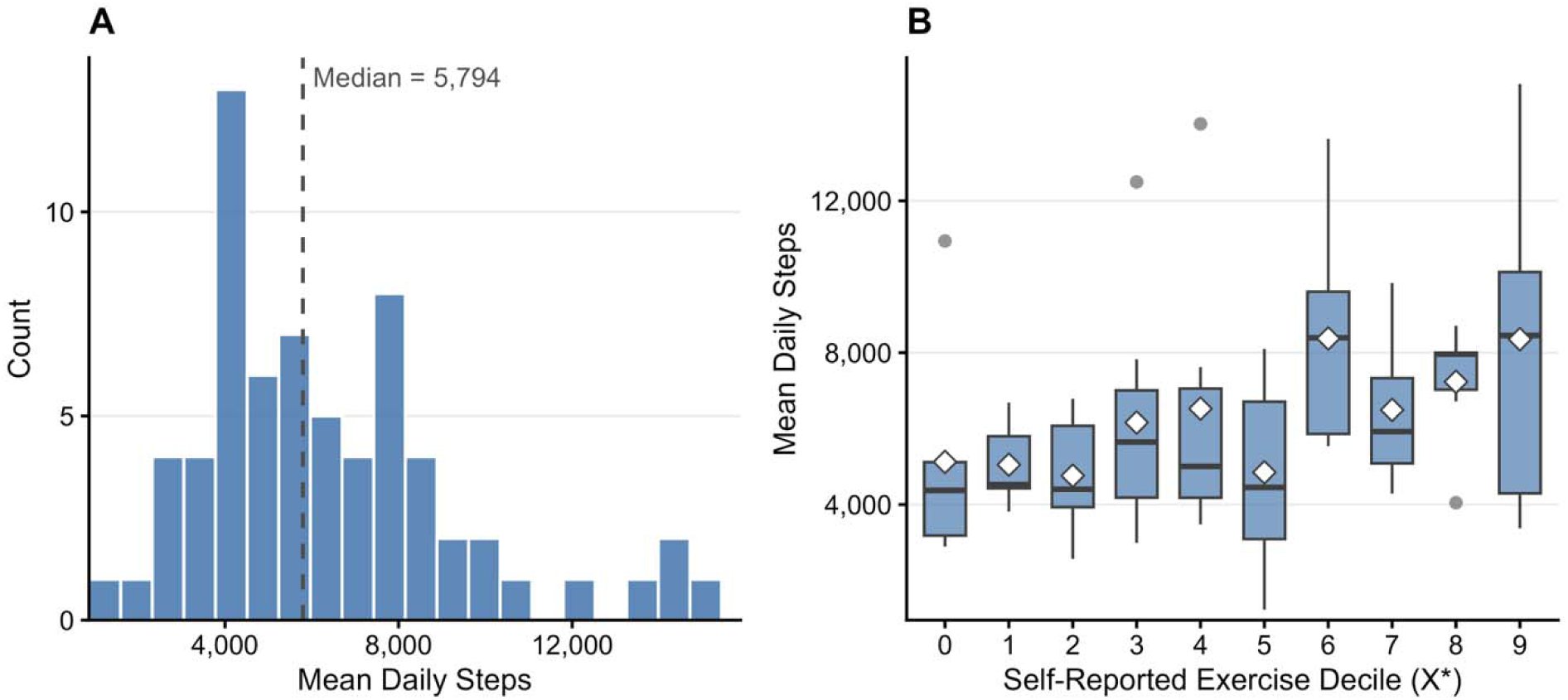
Distribution of Garmin-derived mean daily steps and correspondence with self-reported exercise. Note. (A) Histogram of participant-level mean daily steps (n = 67); dashed line indicates the sample median (5,794 steps/day). (B) Mean daily steps by self-reported exercise decile; boxes show interquartile range and median, whiskers extend to 1.5 × interquartile range, and diamonds indicate decile means.

Sensitivity analyses comparing the primary ≥10-hour valid-day threshold to the more lenient ≥6-hour threshold yielded identical participant-level step-count estimates. The median number of days lost under the stricter criterion was 0 (range: 0-2), and mean daily steps, within-person SD, and CV were identical across thresholds, confirming that the valid-day definition did not materially influence participant-level step count estimates in this dataset.

### 3.2 Evaluation of Self-Reported Physical Activity Using Regression Calibration

Mean daily steps were right-skewed on the raw scale. Examination of calibration model diagnostics supported log-transformation of step counts for better fit, and a linear specification of the calibration model was retained after quadratic and cubic terms for exercise decile were non-significant (details in Supplement S1.3). The calibration model confirmed a statistically significant positive relationship between exercise decile and log-transformed mean daily steps in the linear model (α_₁_ = 0.06, SE = 0.02, p = 0.008), corresponding to an approximate 6.3% increase in mean daily steps per one-decile increase in self-reported exercise. After comparing the measurement approaches for all 16 outcome measures, contrast between approaches was most pronounced for cardiorespiratory fitness. For relative peak V□O_2_, the naive self-report model produced a non-significant coefficient near zero (β = 0.396, 95% CI: −0.008, 0.800; p = 0.054), while the reference Garmin model identified a strong positive association (β = 5.315, 95% CI: 3.309, 7.322; p < 0.001) (**Table 3**). For reference, this coefficient implies that doubling daily step counts from 1,000 to 2,000 is associated with an increase of ∼3.7 mL/kg/min in relative peak V□O_2_, whereas increasing from 4,000 to 5,000 steps corresponds to a smaller increase of ∼1.2 mL/kg/min. Across all 16 outcomes, the self-report coefficient was closer to zero than the corresponding Garmin coefficient. This pattern is consistent with attenuation under a classical measurement error model, and motivated our use of regression calibration to correct for this error structure. The regression-calibrated estimate for relative peak V□O_2_ recovered a directionally consistent and substantially larger effect (β = 7.025, 95% CI: −1.134, 14.249). This coefficient implies that doubling daily step counts (e.g., 1000 to 2000) is associated with an increase of ∼4.9 units in relative peak V□O2, whereas increasing from 4000 to 5000 steps corresponds to a smaller increase of ∼1.6 units. In this model, however, bootstrap standard errors were wide, reflecting uncertainty propagated from the calibration model, and the corrected estimate did not individually reach statistical significance. Comparisons of results for all three models are reported in **Supplementary Table S2**.

**Table 3.** Illustrative three-model comparison: cardiorespiratory fitness outcomes.

| Outcome | Self-report (naive) |  | Observed Garmin |  | RC-corrected |  |
| --- | --- | --- | --- | --- | --- | --- |
|  | Estimate (95% CI) | p | Estimate (95% CI) | p | Estimate (95% CI) □ | p |
| Absolute peak $\dot{V}\text{O}_2$ (L/min) | 0.028<br>(-0.001, 0.057) | 0.060 | 0.292<br>(0.135, 0.449) | <0.001 | 0.491<br>(-0.021, 1.002) | 0.064 |
| Relative peak $\dot{V}\text{O}_2$ (mL/kg/min) | 0.396<br>(-0.008, 0.800) | 0.054 | 5.315<br>(3.309, 7.322) | <0.001 | 7.025<br>(-1.134, 14.249) | 0.071 |

Note. Self-report (naive) predictor: total weekly exercise minutes ranked into deciles; estimate is the coefficient per decile unit. Observed Garmin predictor: log-transformed mean daily step count; estimate is the coefficient per log-step unit. All models adjusted for age, sex, and treatment group. n varies due to missing data. $\dot{V}\text{O}_2$ , oxygen consumption
□ RC-corrected 95% CIs reflect bootstrap uncertainty (B = 2,000, percentile method). RC-corrected p-values are derived from bootstrap standard errors; no RC-corrected estimate reached statistical significance after accounting for calibration uncertainty.

## 4. Discussion

This paper presented a transparent, reproducible SOP for processing commercially available Garmin wearable step-count data into participant-level analytic variables and demonstrated its application in the STRRIDE-PD Reunion study. Beyond validating the pipeline’s technical performance, the application revealed a consistent and clinically meaningful pattern: device-measured daily steps were significantly associated with multiple cardiometabolic and fitness outcomes, while self-reported exercise showed no significant associations with any outcome. A regression calibration analysis formally quantified the attenuation underlying this discrepancy, confirming that the pipeline-derived step counts not only answered the clinical questions of interest but also exposed and quantified a fundamental limitation of the self-report instrument that had previously gone unmeasured.

### 4.1 Step-Count SOP

The processing pipeline produced stable participant-level step-count estimates, with derived summaries identical across both ≥10-hour and ≥6-hour wear-time threshold specifications, reflecting high wear compliance in this cohort. Because the core workflow does not depend on Garmin-specific data structures, the pipeline can be adapted to other consumer devices providing epoch-level timestamped step counts. Consumer-grade wearables have demonstrated good-to-excellent step-count validity across device types (Evenson and others 2015; Evenson and Spade 2020; Fuller and others 2020), including in community-dwelling older adults (Paul and others 2015), supporting their use as a foundation for research pipelines of this kind. Investigators doing so are encouraged to document parameter choices transparently and conduct sensitivity analyses across candidate thresholds, as illustrated here, to verify that conclusions are robust to the specific processing decisions made. In populations with lower wear compliance than observed in this study, such as older adults with cognitive impairment or significant mobility limitations, the wear-time inference rules may need to be relaxed or supplemented with diary-based wear logs to avoid systematically excluding the least active participants and introducing selection bias into derived estimates.

Wearable-derived mean daily steps were significantly associated with 9 of 16 reunion biometric outcomes. This pattern is consistent with the growing literature linking higher daily step counts to improved cardiometabolic and fitness markers (Kraus and others 2019a; Paluch and others 2022; Saint-Maurice and others 2020; Tudor-Locke and others 2011). Notably, the sample median of 5,794 steps/day falls within the range at which health benefits begin to plateau in older adults (approximately 6,000-8,000 steps/day) (Paluch and others 2022; Tudor-Locke and others 2011), suggesting the associations detected here reflect PA levels within a clinically meaningful and modifiable range for this cohort. The pattern of associations is consistent with the expected biology: significant findings clustered among outcomes reflecting total daily energy expenditure, including body composition, waist circumference, and cardiorespiratory fitness, while outcomes more sensitive to dietary factors or medication use, such as low-density lipoprotein cholesterol (LDL-C), triglycerides, and glucose showed no significant associations. These null findings should not be interpreted as evidence of no effect. Rather, limited statistical power with a sample size of 67 participants and the relatively short monitoring window likely constrained the ability to detect associations of plausible magnitude for these outcomes.

Among the significant associations detected, cardiorespiratory fitness warrants particular attention, as cardiorespiratory fitness is an independent predictor of cardiovascular disease risk, all-cause mortality, and functional dependence (Ross and others 2016). Meta-analytic evidence indicates that each one metabolic equivalent of task (MET; 1 MET ≈ 3.5 mL/kg/min) higher level of cardiorespiratory fitness is associated with a 13-15% reduction in all-cause mortality and cardiovascular events (Kodama and others 2009). Doubling daily steps from approximately 1,000 to 2,000 steps/day corresponds to an estimated increase of approximately 1 MET in relative peak V□O_2_ – a difference reaching the threshold associated with meaningful reductions in mortality risk (Kodama and others 2009). An equivalent absolute step increase at a higher baseline – from 4,000 to 5,000 steps/day – corresponds to a smaller, though still meaningful gain (∼1.2 mL/kg/min). This pattern is consistent with the curvilinear dose-response relationship between PA and health outcomes in which the greatest marginal benefits accrue among the least active individuals (Kraus and others 2019b). These findings demonstrate that wearable-derived step-count data, processed through a reproducible pipeline, can detect PA-fitness associations of clinically meaningful magnitude in aging cohorts – associations that the self-reported PA measure failed to capture.

### 4.2 Self-Reported Physical Activity

The pipeline-derived data allows us to highlight two compounding and distinct limitations of the self-report instrument. The first is a construct mismatch where the questionnaire captures only intentional structured exercise, not total daily movement. The questionnaire therefore captures a fundamentally narrower behavioral construct than wearable-derived step counts(Dyrstad and others 2014; Prince and others 2020). This distinction is particularly consequential in older adults, for whom light-intensity incidental activity constitutes a meaningful share of daily energy expenditure and carries independent health benefits (LaMonte and others 2017). The second is classical measurement error. Even within the targeted construct, the crude decile ranking introduces noise that attenuates associations toward the null (Adams and others 2005; Prince and others 2008).

Because these two sources of error are intertwined, regression calibration cannot cleanly isolate and correct for one while leaving the other untouched (Carroll and Carroll 2006; Keogh and others 2020; Rosner and others 1989). As illustrated by the relative peak V□O_2_ example, RC-corrected estimates do not simply recover to the magnitude of the reference Garmin coefficients. This suggests the calibration model is partly compensating for the construct gap in addition to the noise introduced by the decile ranking. Nonetheless, RC meaningfully shifted the self-report estimate toward the Garmin-observed magnitude (from near zero (β = 0.396) to β = 7.025) indicating that the near-null self-report association reflects measurement artifact rather than a true absence of effect, even though bootstrap uncertainty in this sample precluded statistical significance. Had this study relied solely on questionnaire data, as many aging cohort studies with limited resources must, the conclusion would have been that PA was unrelated to fitness and cardiometabolic health at the Reunion timepoint. This conclusion is an artifact of measurement rather than a biological reality.

This application represents a somewhat unusual case for regression calibration. Because Garmin data were available for all analytic participants, predicted log step counts were not strictly necessary to estimate associations at the Reunion timepoint. In a more typical setting, the objective reference measure would be available only for a subset of participants, and the calibration model would be used to impute predicted values for those without it. The present analysis is therefore best understood as a proof of concept, confirming that the calibration framework is feasible in this cohort and that self-reported PA introduces meaningful attenuation relative to the objective reference. This has direct implications for future work given many earlier timepoints in aging cohort studies predate wearable device availability. Thus, a calibration model built from concurrent self-report and Garmin data at the Reunion study could be used to reconstruct predicted log step counts at historical timepoints for participants without prior device-based monitoring, enabling longitudinal step count trajectories to be estimated across the full cohort.

### 4.3 Limitations

The most important limitation of this application is the relatively short monitoring window. Garmin data were collected over approximately one week, which may not fully represent participants’ habitual activity levels. Participants were given the device at their first in-person Reunion assessment visit and instructed to wear it continuously; however, days with data may nonetheless correspond disproportionately to days of higher activity if participants were selectively more active during the monitoring window, potentially overestimating typical daily step counts.

A second limitation concerns wear-time inference. Wear time was operationally defined as the interval between the first and last non-zero step epochs within each day. Therefore, a low activity day (*e.g.,* a day on which a participant did not take a step until midday or retired early) may be assigned artificially short wear windows. The present analysis used a flexible, transparent wear-time definition that does not rely on ancillary signals such as device-reported wear logs or heart rate data – an approach that is broadly applicable across consumer device contexts where such metadata are unavailable, and that has been applied in comparable Garmin-based studies in older adults (Colon-Emeric and others 2026). Heart rate data were not incorporated into wear-time estimation in this application. Future work could explore integrating heart rate-based on-body detection to further validate or refine epoch-level wear classification, and such data are available in this cohort for that purpose.

A third limitation concerns the sensitivity analysis comparing the ≥10-hour and ≥6-hour valid-day thresholds. While participant-level step-count estimates were identical across both thresholds, this similarity largely reflects the high wear compliance of this sample with a median of zero days lost under the stricter criterion, rather than a rigorous stress-test of the pipeline. Future work should examine stricter thresholds, such as ≥16 hours, which better align with conventions in the 24-hour wear protocol literature, to characterize whether step-count estimates remain stable in samples with more variable compliance.

Finally, the small analytic sample of 67 participants limited the calibration model precision. The bootstrap confidence intervals were wide for all corrected estimates, reflecting the substantial uncertainty propagated from the calibration model into the outcome models.

## 5. Conclusions

This work demonstrated that a transparent, reproducible processing pipeline for commercial wearable data can serve as more than a data cleaning tool – the pipeline enables rigorous inference about PA and health. Consumer-device measured step counts detected significant associations with cardiometabolic and fitness outcomes that were entirely undetectable using self-reported PA. Moreover, the regression calibration framework formally quantified the attenuation responsible for that failure. Together, these findings underscore that measurement approach is not a peripheral analytic detail, but a determinant of the scientific conclusions drawn about PA and health in aging populations. The pipeline developed here provides a practical and transferable foundation for investigators seeking to maximize the inferential value of wearable data in longitudinal research settings.

## ACKNOWLEDGEMENTS

We would like to thank all of the STRRIDE-PD Reunion participants, staff members, and student interns.

## AUTHOR CONTRIBUTIONS

Nannan Bo: conceptualization, data curation, formal analysis, methodology, visualization, writing – original draft, writing – review and editing. Alyssa Sudnick: data curation, formal analysis, investigation, project administration, writing – review and editing. Julie Counts: investigation, project administration, writing – review and editing. Katie Kennedy: investigation, writing – review and editing. Agustin Saldana: investigation, writing – review and editing. Katherine Collins-Bennett: writing – review and editing.

William Bennett: investigation, writing – review and editing. Johanna Johnson: project administration, writing – review and editing. Kim Huffman: writing – review and editing. Amanda Paluch: writing – review and editing. Marissa Ashner: methodology, writing – review and editing. William Kraus: conceptualization, funding acquisition, resources, supervision, writing – review and editing. Sarah Peskoe: conceptualization, formal analysis, methodology, supervision, writing – review and editing. Leanna Ross: conceptualization, data curation, investigation, project administration, supervision, writing – review and editing.

## DATA AVAILABILITY

The raw data supporting the conclusions of this article will be made available by the authors without undue reservation. Requests to access the data should be directed to WEK.

## DECLARATION OF GENERATIVE AI IN THE MANUSCRIPT PREPARATION PROCESS

During the preparation of this work, the authors used Claude (Anthropic) to support language editing and content organization. After using this tool/service, the authors reviewed and edited the content as needed and take full responsibility for the content of the published article.

## Supplement

### S1. Methods: Regression Calibration Framework

#### S1.1 Measurement error framework and notation

The regression calibration framework follows the guidance of Keogh et al. (2020). The framework is designed for settings where the true exposure is unobserved and only an error-prone surrogate X* is available. Notation in the context of this paper is as follows:

X^*^ = self-reported exercise decile (the error-prone measure)

log (X) = Garmin-derived log mean daily steps (the imperfect reference measure)

z = vector of exactly measured covariates (age, sex, treatment group)

Y = reunion biometric outcome

ε ∼ N(0, σ²) = the residual error term

The goal of regression calibration is to obtain a corrected estimate of the association between objective step-count-indexed PA and each outcome Y by replacing *X*^*^ with the calibration-predicted value (log (*X*) | *X*^*^,*z*) in the outcome model. Because Garmin step counts serve as an imperfect rather than gold-standard reference, corrected estimates represent associations with Garmin-indexed activity during the monitoring window, adjusted for attenuation from the crude decile ranking.

#### S1.2 Model assumptions

##### Classical measurement error structure

The self-reported exercise decile X* is assumed to approximate the objective reference X plus a random error term with mean zero that is independent of X. This classical additive error structure was selected for three reasons (Carroll and Carroll 2006; Keogh and others 2020). First, the decile’s construction as a rank-based summary of categorical self-report items is consistent with noise that is roughly symmetric and mean-zero across participants, without a strong a priori expectation of directional bias. Second, classical ME is the foundational framework recommended by the STRATOS guidance for settings where a single error-prone measure and an internal reference are available and where the primary goal is attenuation correction in linear outcome models (Keogh and others 2020). Third, the empirical pattern of results — in which the self-report coefficient was closer to zero than the observed Garmin coefficient for every one of the 16 outcomes examined — is consistent with the attenuation predicted under classical ME and supports the model choice post hoc. It is worth noting, however, that because the calibration model predicts log-transformed Garmin steps rather than steps on the raw scale, attenuation of the naive self-report coefficients toward zero is not analytically guaranteed by the classical ME framework alone; the consistent attenuation pattern observed here is therefore an empirical finding that reinforces, rather than follows automatically from, the model assumption. Systematic underreporting in specific subgroups (e.g., by sex or treatment arm) cannot be ruled out and would violate the mean-zero error assumption; this limitation should be acknowledged when interpreting corrected estimates.

##### Non-differential measurement error

The error in X* is assumed to be independent of the outcome Y conditional on true exposure X. This is plausible for the biometric outcomes collected at the reunion clinic visit, where measurements were obtained by trained staff independently of how participants characterized their exercise habits.

##### Linearity of the calibration model

The relationship between exercise decile and log-transformed mean daily steps is assumed to be approximately linear across the observed range of the decile (0–9). Visual inspection of mean daily steps across deciles supported a broadly monotone increasing pattern without evidence of strong curvature (Figure 2B). To further evaluate this assumption, we examined whether higher-order polynomial terms for exercise decile improved model fit in the calibration model. Neither the quadratic nor cubic terms were statistically significant (p = 0.903 and p = 0.846, respectively), and model fit did not improve meaningfully with their inclusion. The linear specification was therefore retained as the primary calibration model.

##### Homoscedasticity of calibration residuals

The variance of the error in the calibration equation is assumed to be constant across the range of the predictor. This assumption was evaluated using the Breusch-Pagan test (Breusch and Pagan 1979) and residual diagnostic plots. On the raw step-count scale, the Breusch-Pagan test indicated heteroscedasticity (p = 0.246), while on the log scale residuals were substantially more homoscedastic (p = 0.560.

#### S1.3 Log-Transformation of the Reference Measure

Garmin step counts were right-skewed on the raw scale, motivating consideration of a log-transformation. We compared simple calibration regressions on both scales using R², residual standard error, and the Breusch-Pagan test for heteroscedasticity. Neither specification yielded a statistically significant Breusch-Pagan test (raw scale: p = 0.246; log scale: p = 0.560), and R² was similar across both (0.135 raw vs. 0.126 log). The decision to use the log scale therefore rested on residual diagnostic plots, which indicated more consistent variance on the log scale than on the raw scale (Figure S1), consistent with the recommendation in Keogh et al. (2020) to prefer the scale on which homoscedasticity is better supported even when formal tests do not reach significance. All subsequent calibration and outcome models used log-transformed mean daily steps as the reference measure.

#### S1.4 Calibration Model Specification

The calibration model estimated (log (*X*) |*X*^*^, *z*) by regressing log-transformed mean daily Garmin steps on exercise decile and covariates:

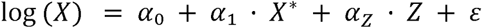

where Z includes age (continuous), sex (men vs. women), and treatment group (three indicators, reference = Low/Moderate arm). Covariates were included in the calibration equation to ensure that the predicted values condition on the same information used in the outcome models.

Model diagnostics included residual plots, the Breusch-Pagan test, and Cook’s distance to identify influential observations. Four observations exceeded the 4/n Cook’s distance threshold; these were retained in the primary analysis given the small sample size and the absence of evidence for systematic deviation from the model.

#### S1.5 Naive and Reference Outcome Models

Prior to applying regression calibration, two sets of outcome models were fit to provide empirical anchors for the RC-corrected estimates.

The naive outcome model for each biometric outcome Y was:

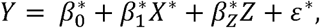

where 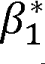 is the naive estimate of the association between self-reported PA and the outcome, which is expected to be attenuated toward zero relative to the true PA-outcome association due to measurement error in *X*^*^.

The reference outcome model for each outcome Y was:

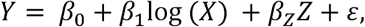

where β_1_ reflects the association between objectively measured PA and the outcome, serving as a benchmark against which the naive and RC-corrected estimates are compared.

Naive estimates reflect the degree of attenuation induced by measurement error in *X*^*^, while reference estimates reflect what associations look like when the better-measured variable is used directly, without calibration correction. We compared the naive, reference, and RC-corrected model specifications to quantify the inferential consequences of measurement approach.

#### S1.6 Regression Calibration Outcome Models

Regression calibration outcome models substituted the calibration-predicted value 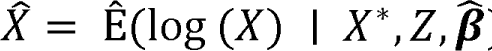) for the raw exercise decile in each of the 16 outcome models. The RC-corrected predictor 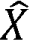 is expressed on the same scale as the reference measure (log-transformed mean daily step counts) because the calibration model predicts log (*X*) from *X*^*^ and *z*. The same structure of the outcome model was fit:

The naive outcome model for each biometric outcome Y was:

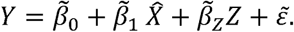

Consequently, RC-corrected coefficients are directly comparable to those from the observed Garmin reference model: a one-unit increase in 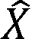 corresponds to a one-log-step increase in predicted PA.

The correction factor for each outcome is the ratio of the RC-corrected to the naive estimate, quantifying the degree of attenuation in the naive model. Because regression calibration applies a single scalar correction derived from the calibration model slope (â_₁_) and the variance structure of *X*^*^, the correction factor is constant across all outcomes at 16.46. A correction factor of this magnitude is mechanistically consistent with a calibration R² of 0.21: when the error-prone measure explains only 21% of the variance in the reference, the adjustment required to recover the attenuated signal must be correspondingly large.

#### S1.7 Bootstrap Variance Estimation

Naive standard errors from the regression calibration procedure underestimate true uncertainty because calibration-predicted values are treated as known constants rather than estimates with their own sampling variability (Buonaccorsi 2010; Carroll and Carroll 2006). To obtain valid standard errors and confidence intervals, a non-parametric bootstrap was implemented across the full estimation pipeline (B = 2,000; seed = 2025). In each bootstrap iteration, a sample of 67 participants was drawn with replacement, the calibration model was refit, calibration-predicted values were generated, and the outcome model was fit using these predicted values. Bootstrap standard errors were computed as the standard deviation of the bootstrap distribution of each RC coefficient, and 95% bootstrap percentile confidence intervals were constructed from the 2.5th and 97.5th percentiles. Bootstrap-based p-values were derived from these standard errors. Bootstrap standard errors were substantially larger than model-based standard errors for all 16 outcomes, and no RC-corrected estimate reached statistical significance after accounting for calibration uncertainty.

### S2. Health-Related Outcomes

The present analysis examined associations between physical activity (PA) and 16 health-related outcomes assessed at the STRRIDE-PD Reunion timepoint, spanning five domains: anthropometric, body composition, cardiorespiratory fitness, blood-based cardiometabolic, and hemodynamic measures. Outcomes were selected to cover the breadth of cardiometabolic and fitness constructs assessed in the reunion protocol and were analyzed as continuous variables without transformation. Table S1 provides the full list of outcomes, their units, and a brief description of the assessment method or data source for each.

### S3. Primary Regression Results: Three-Model Comparison

Table S2 presents the full three-model comparison: naive self-report, observed Garmin, and regression-calibrated (RC), across all 16 reunion outcomes. For each outcome, the naive model used the raw self-reported exercise decile as the PA predictor; the reference model used log-transformed mean daily Garmin steps; and the RC model substituted the calibration-predicted value from the primary calibration model. All models adjusted for age, sex, and treatment group. Analytic 95% confidence intervals are reported for the naive and observed Garmin models; bootstrap percentile confidence intervals (B = 2,000, seed = 2025) are reported for the RC-corrected estimates.

The self-report model produced uniformly small, non-significant coefficients across all 16 outcomes. The observed Garmin model identified statistically significant associations for 9 of 16 outcomes spanning body composition, lipid profiles, and cardiorespiratory fitness. RC-corrected estimates were directionally consistent with the naive and reference estimates for all 16 outcomes, with no sign reversals; however, no RC-corrected estimate reached statistical significance after bootstrap variance estimation, reflecting the substantial uncertainty propagated from the calibration model into the corrected estimates.

**Table S1.**
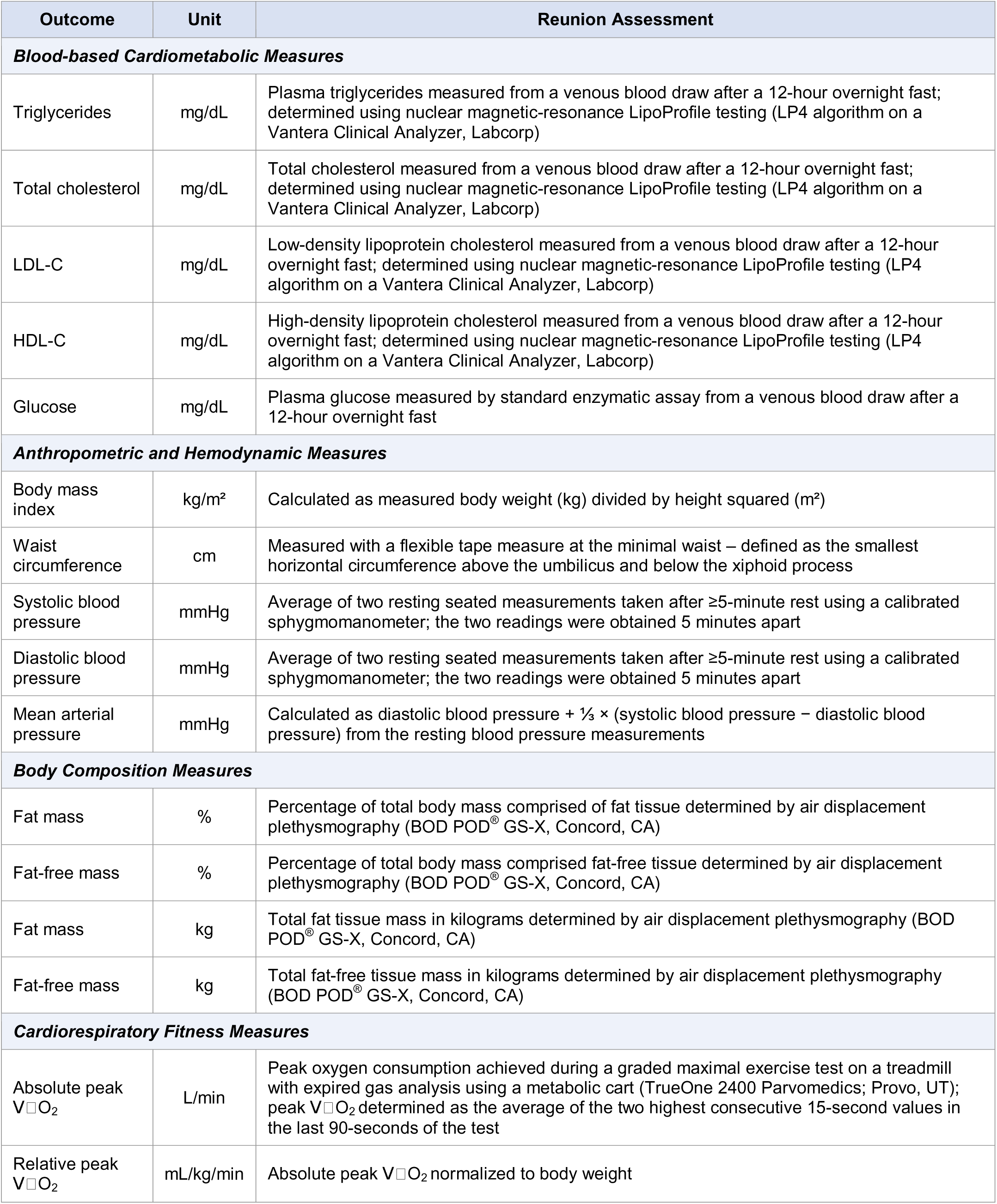
Health-related outcomes assessed in the STRRIDE-PD Reunion study included in the physical activity analysis.

| Outcome | Unit | Reunion Assessment |
| --- | --- | --- |
| **Blood-based Cardiometabolic Measures** |  |  |
| Triglycerides | mg/dL | Plasma triglycerides measured from a venous blood draw after a 12-hour overnight fast; determined using nuclear magnetic-resonance LipoProfile testing (LP4 algorithm on a Vantera Clinical Analyzer, Labcorp) |
| Total cholesterol | mg/dL | Total cholesterol measured from a venous blood draw after a 12-hour overnight fast; determined using nuclear magnetic-resonance LipoProfile testing (LP4 algorithm on a Vantera Clinical Analyzer, Labcorp) |
| LDL-C | mg/dL | Low-density lipoprotein cholesterol measured from a venous blood draw after a 12-hour overnight fast; determined using nuclear magnetic-resonance LipoProfile testing (LP4 algorithm on a Vantera Clinical Analyzer, Labcorp) |
| HDL-C | mg/dL | High-density lipoprotein cholesterol measured from a venous blood draw after a 12-hour overnight fast; determined using nuclear magnetic-resonance LipoProfile testing (LP4 algorithm on a Vantera Clinical Analyzer, Labcorp) |
| Glucose | mg/dL | Plasma glucose measured by standard enzymatic assay from a venous blood draw after a 12-hour overnight fast |
| **Anthropometric and Hemodynamic Measures** |  |  |
| Body mass index | kg/m<sup>2</sup> | Calculated as measured body weight (kg) divided by height squared (m<sup>2</sup>) |
| Waist circumference | cm | Measured with a flexible tape measure at the minimal waist – defined as the smallest horizontal circumference above the umbilicus and below the xiphoid process |
| Systolic blood pressure | mmHg | Average of two resting seated measurements taken after ≥5-minute rest using a calibrated sphygmomanometer; the two readings were obtained 5 minutes apart |
| Diastolic blood pressure | mmHg | Average of two resting seated measurements taken after ≥5-minute rest using a calibrated sphygmomanometer; the two readings were obtained 5 minutes apart |
| Mean arterial pressure | mmHg | Calculated as diastolic blood pressure + $\frac{1}{3} \times$ (systolic blood pressure – diastolic blood pressure) from the resting blood pressure measurements |
| **Body Composition Measures** |  |  |
| Fat mass | % | Percentage of total body mass comprised of fat tissue determined by air displacement plethysmography (BOD POD<sup>®</sup> GS-X, Concord, CA) |
| Fat-free mass | % | Percentage of total body mass comprised fat-free tissue determined by air displacement plethysmography (BOD POD<sup>®</sup> GS-X, Concord, CA) |
| Fat mass | kg | Total fat tissue mass in kilograms determined by air displacement plethysmography (BOD POD<sup>®</sup> GS-X, Concord, CA) |
| Fat-free mass | kg | Total fat-free tissue mass in kilograms determined by air displacement plethysmography (BOD POD<sup>®</sup> GS-X, Concord, CA) |
| **Cardiorespiratory Fitness Measures** |  |  |
| Absolute peak $\dot{V}\text{O}_2$ | L/min | Peak oxygen consumption achieved during a graded maximal exercise test on a treadmill with expired gas analysis using a metabolic cart (TrueOne 2400 Parvomedics; Provo, UT); peak $\dot{V}\text{O}_2$ determined as the average of the two highest consecutive 15-second values in the last 90-seconds of the test |
| Relative peak $\dot{V}\text{O}_2$ | mL/kg/min | Absolute peak $\dot{V}\text{O}_2$ normalized to body weight |

**Table S2.**
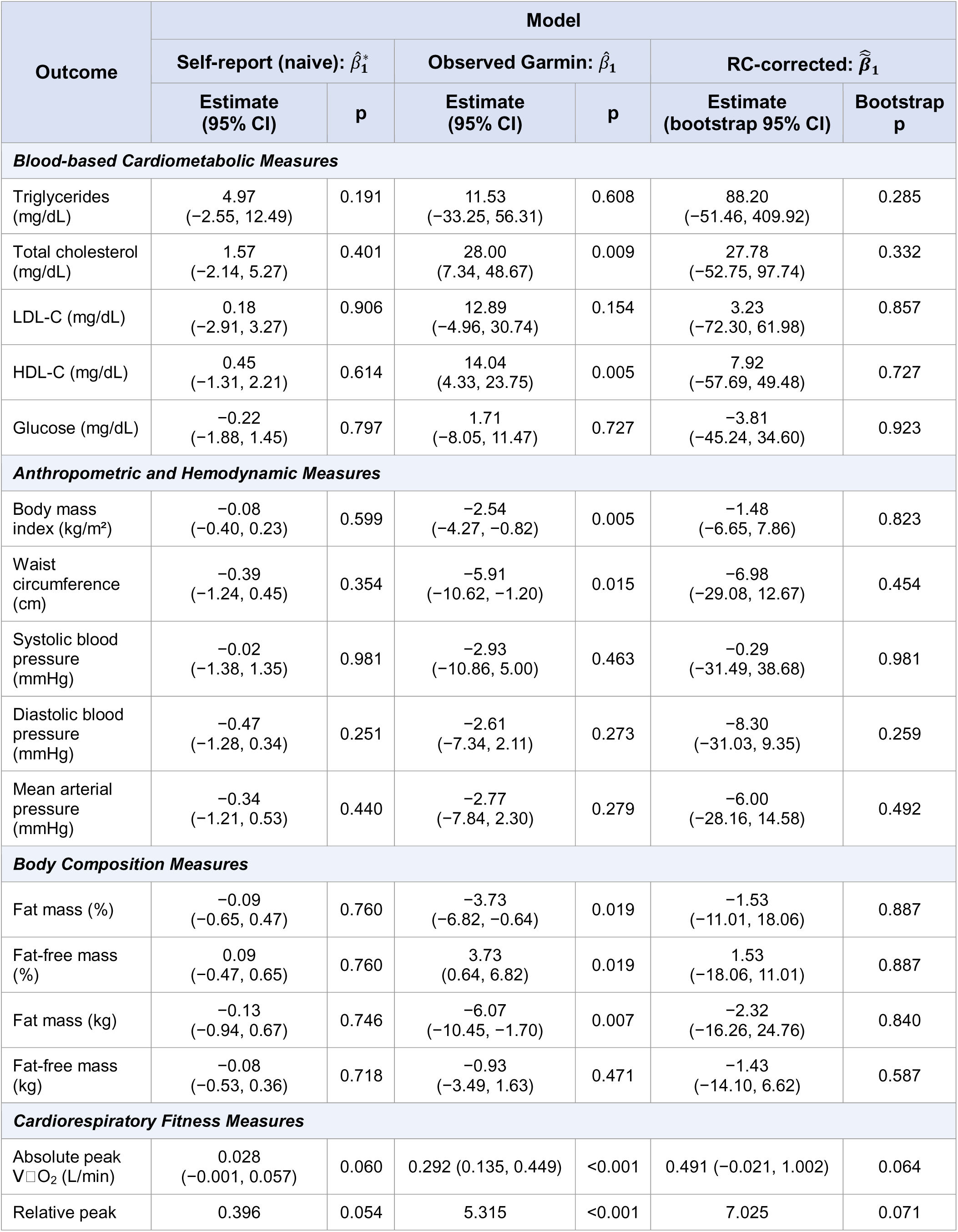
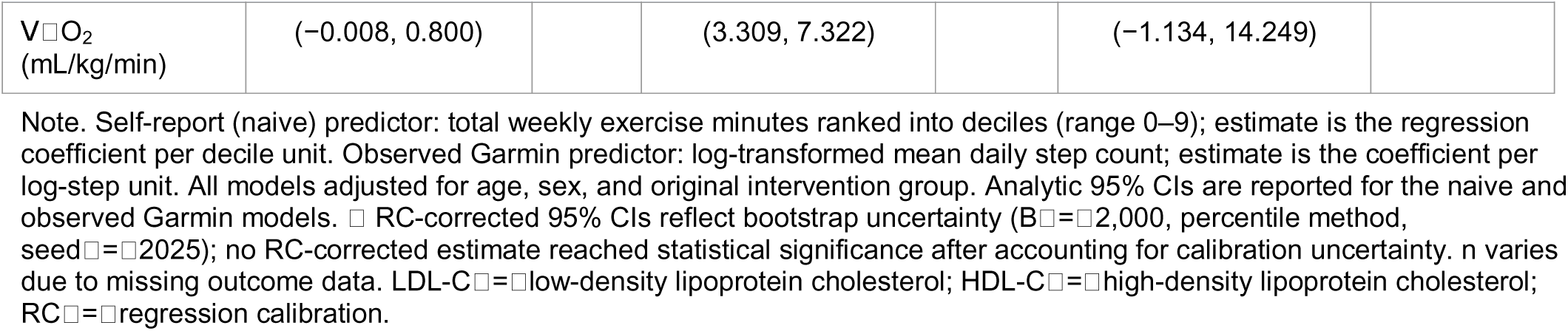
Three-model comparison: naive self-report, observed Garmin, and RC-corrected estimates with bootstrap uncertainty.

| Outcome | Model |  |  |  |  |  |
| --- | --- | --- | --- | --- | --- | --- |
| | Self-report (naive): $\hat{\beta}_1^*$ | | Observed Garmin: $\hat{\beta}_1$ | | RC-corrected: $\hat{\hat{\beta}}_1$ | |
|  | Estimate<br>(95% CI) | p | Estimate<br>(95% CI) | p | Estimate<br>(bootstrap 95% CI) | Bootstrap<br>p |
| **Blood-based Cardiometabolic Measures** |  |  |  |  |  |  |
| Triglycerides (mg/dL) | 4.97<br>(-2.55, 12.49) | 0.191 | 11.53<br>(-33.25, 56.31) | 0.608 | 88.20<br>(-51.46, 409.92) | 0.285 |
| Total cholesterol (mg/dL) | 1.57<br>(-2.14, 5.27) | 0.401 | 28.00<br>(7.34, 48.67) | 0.009 | 27.78<br>(-52.75, 97.74) | 0.332 |
| LDL-C (mg/dL) | 0.18<br>(-2.91, 3.27) | 0.906 | 12.89<br>(-4.96, 30.74) | 0.154 | 3.23<br>(-72.30, 61.98) | 0.857 |
| HDL-C (mg/dL) | 0.45<br>(-1.31, 2.21) | 0.614 | 14.04<br>(4.33, 23.75) | 0.005 | 7.92<br>(-57.69, 49.48) | 0.727 |
| Glucose (mg/dL) | -0.22<br>(-1.88, 1.45) | 0.797 | 1.71<br>(-8.05, 11.47) | 0.727 | -3.81<br>(-45.24, 34.60) | 0.923 |
| **Anthropometric and Hemodynamic Measures** |  |  |  |  |  |  |
| Body mass index (kg/m<sup>2</sup>) | -0.08<br>(-0.40, 0.23) | 0.599 | -2.54<br>(-4.27, -0.82) | 0.005 | -1.48<br>(-6.65, 7.86) | 0.823 |
| Waist circumference (cm) | -0.39<br>(-1.24, 0.45) | 0.354 | -5.91<br>(-10.62, -1.20) | 0.015 | -6.98<br>(-29.08, 12.67) | 0.454 |
| Systolic blood pressure (mmHg) | -0.02<br>(-1.38, 1.35) | 0.981 | -2.93<br>(-10.86, 5.00) | 0.463 | -0.29<br>(-31.49, 38.68) | 0.981 |
| Diastolic blood pressure (mmHg) | -0.47<br>(-1.28, 0.34) | 0.251 | -2.61<br>(-7.34, 2.11) | 0.273 | -8.30<br>(-31.03, 9.35) | 0.259 |
| Mean arterial pressure (mmHg) | -0.34<br>(-1.21, 0.53) | 0.440 | -2.77<br>(-7.84, 2.30) | 0.279 | -6.00<br>(-28.16, 14.58) | 0.492 |
| **Body Composition Measures** |  |  |  |  |  |  |
| Fat mass (%) | -0.09<br>(-0.65, 0.47) | 0.760 | -3.73<br>(-6.82, -0.64) | 0.019 | -1.53<br>(-11.01, 18.06) | 0.887 |
| Fat-free mass (%) | 0.09<br>(-0.47, 0.65) | 0.760 | 3.73<br>(0.64, 6.82) | 0.019 | 1.53<br>(-18.06, 11.01) | 0.887 |
| Fat mass (kg) | -0.13<br>(-0.94, 0.67) | 0.746 | -6.07<br>(-10.45, -1.70) | 0.007 | -2.32<br>(-16.26, 24.76) | 0.840 |
| Fat-free mass (kg) | -0.08<br>(-0.53, 0.36) | 0.718 | -0.93<br>(-3.49, 1.63) | 0.471 | -1.43<br>(-14.10, 6.62) | 0.587 |
| **Cardiorespiratory Fitness Measures** |  |  |  |  |  |  |
| Absolute peak $\dot{V}\text{O}_2$ (L/min) | 0.028<br>(-0.001, 0.057) | 0.060 | 0.292 (0.135, 0.449) | <0.001 | 0.491 (-0.021, 1.002) | 0.064 |
| Relative peak | 0.396 | 0.054 | 5.315 | <0.001 | 7.025 | 0.071 |
| | Self-report (naive): $\hat{\beta}_1^*$ | | Observed Garmin: $\hat{\beta}_1$ | | RC-corrected: $\hat{\hat{\beta}}_1$ | |
|  | Estimate<br>(95% CI) | p | Estimate<br>(95% CI) | p | Estimate<br>(bootstrap 95% CI) | Bootstrap<br>p |
| $\dot{V}\text{O}_2$<br>(mL/kg/min) | (−0.008, 0.800) | | (3.309, 7.322) | | (−1.134, 14.249) | |

Note. Self-report (naive) predictor: total weekly exercise minutes ranked into deciles (range 0–9); estimate is the regression coefficient per decile unit. Observed Garmin predictor: log-transformed mean daily step count; estimate is the coefficient per log-step unit. All models adjusted for age, sex, and original intervention group. Analytic 95% CIs are reported for the naive and observed Garmin models. RC-corrected 95% CIs reflect bootstrap uncertainty ( $B=2,000$ , percentile method, seed=2025); no RC-corrected estimate reached statistical significance after accounting for calibration uncertainty. n varies due to missing outcome data. LDL-C=low-density lipoprotein cholesterol; HDL-C=high-density lipoprotein cholesterol; RC=regression calibration.

**Figure S1.**
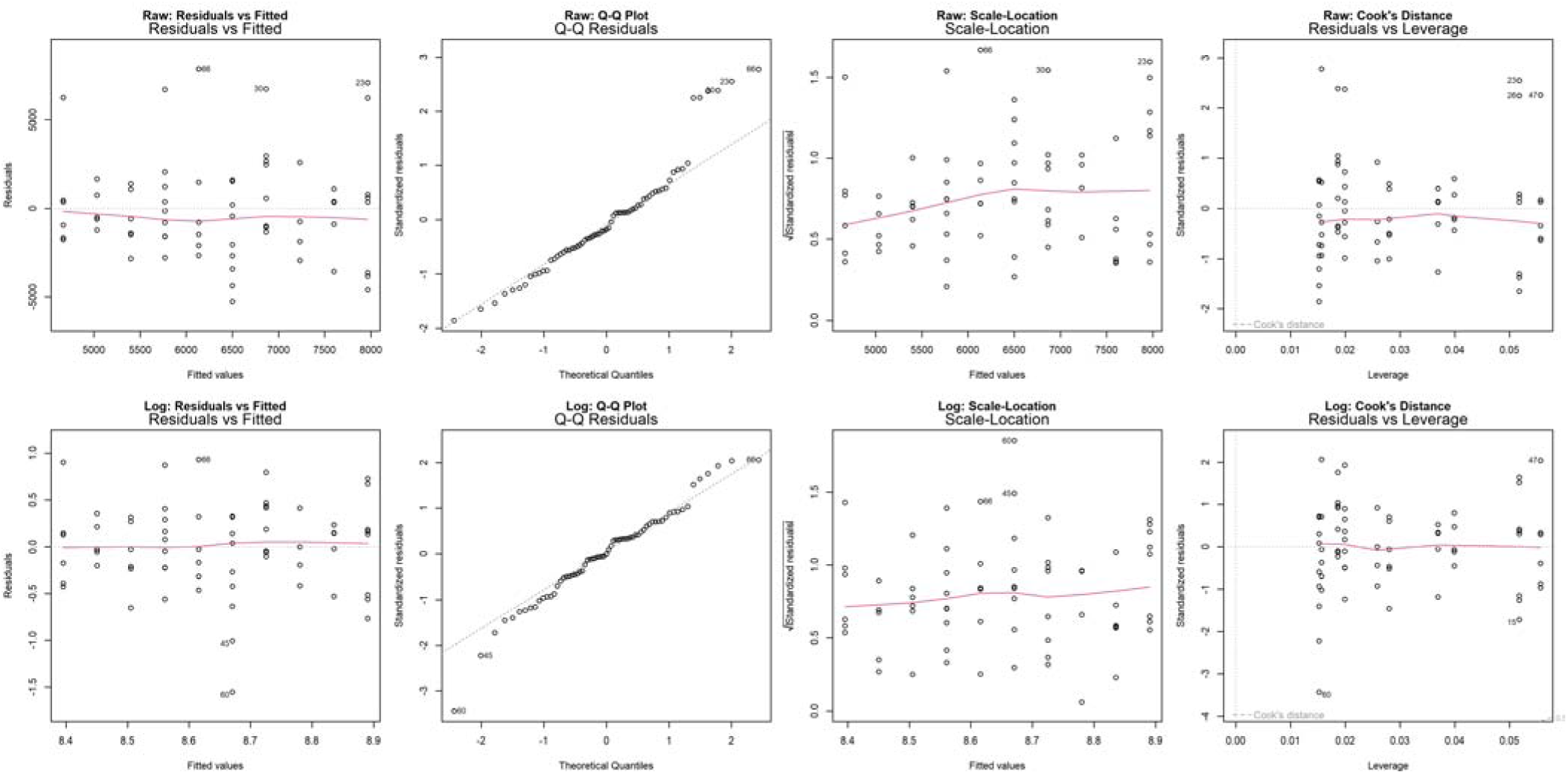
Calibration model diagnostic plots: raw scale (top) vs. log scale (bottom). Note. Each panel shows four standard regression diagnostics for the simple calibration regression of Garmin step counts on self-reported exercise decile (without covariates). On the raw scale, the scale-location plot shows a rising trend in standardized residual variance across fitted values, and the Q-Q plot shows upper-tail deviation consistent with residual right-skew. On the log scale, the residuals vs. fitted and scale-location plots show flat loess lines and more uniform residual spread across the range of fitted values, supporting the homoscedasticity assumption. The Q-Q plot indicates acceptable normality on the log scale, with mild lower-tail deviation attributable to two low-activity participants (observations 61 and 16), who also appear as high-leverage points in the residuals vs. leverage plot. No observations exceed Cook’s distance contours on the log scale.

